# A Low-Cost, Microcontroller-Based Gas Delivery System for Respiratory Stimuli in MRI Studies

**DOI:** 10.64898/2026.05.06.26351951

**Authors:** Nicholas P. Blockley, Ahlam A. Alzaidi, Colette C. Milbourn, Daniel P. Bulte, Andrew Rudgewick-Brown, Sebastian W. Rieger

## Abstract

**Purpose:** To present the design and validation of a lowlZIcost, microcontrollerlZIbased gas delivery system that automates fixed inspired respiratory stimuli for MRI experiments.

**Methods:** The system uses three solenoid valves controlled by an ArduinolZIbased circuit to switch between premixed medical gases according with predefined timings. By using the MRI scanner external timing signal, gas delivery can be synchronised with image acquisition. The system was constructed using commercially available components, with a material cost ∼£650 GBP (∼$880 USD). The system was integrated with a singlelZIuse breathing circuit and evaluated using hypercapnic and hyperoxic stimulus paradigms. EndlZItidal oxygen (O_2_) and carbon dioxide (CO_2_) were measured using a respiratory gas analyser and physiological responses were assessed using BOLD MRI at 3 T.

**Results:** The system delivered reliable, repeatable gas transitions during MRIlZItriggered protocols. During hypercapnia (elevated carbon dioxide level, 5% CO_2_) (n=15), the mean increase in endlZItidal CO_2_ was 8.7±1.8LJmmHg from a baseline of 32.2±3.1LJmmHg, producing a mean grey matter BOLD signal increase of 3.2±1.7%. During hyperoxia (elevated oxygen level, 60.5% O_2_) (n=15), the mean increase in endlZItidal O_2_ was 292.3±59.0LJmmHg from a baseline of 114.5±10.7LJmmHg, with an associated BOLD signal change of 1.2±1.7%. Across both protocols respiratory and BOLD responses were consistent across participants.

**Conclusion:** This microcontrollerlZIbased system provides an inexpensive and reliable method for administering fixed inspired respiratory stimuli with automated MRI synchronisation. It offers an intermediate option between manual systems and commercial gas blenders, making it well suited for technical and methodological studies in cerebrovascular reactivity, hyperoxialZIBOLD and related applications.

## Introduction

The combination of respiratory stimuli and the blood oxygen level dependent (BOLD) signal provide a powerful tool for investigating many aspects of brain physiology. This includes cerebrovascular reactivity (CVR)^1^, cerebral blood volume (CBV)^2^, cerebral metabolic rate of oxygen consumption (CMRO_2_)^3^ and vessel size imaging (VSI)^4,5^. Critical to these techniques is the delivery of gases with altered carbon dioxide (CO_2_) and/or oxygen (O_2_) concentrations in a predetermined order and timing. Such systems consist of three main components: a gas delivery system to mix or switch between the gas sources, a breathing circuit for the participant to breathe from and a gas analyser to measure changes in expired gases due to the stimulus. Whilst there are several commercial options for the latter two components, ranging in complexity and cost, there are limited options for the gas delivery system.

At one end are systems like the one described by Lu et al.^6^, where pre-filled Douglas bags are connected via a three-way valve to a breathing circuit, allowing the inhaled gas to be switched between the bag and room air. This is known as a fixed inspired challenge (FIC) because the inhaled content of CO_2_ and/or O_2_ is fixed. Its advantages are simplicity and low cost, whilst disadvantages include the need to have a researcher within the magnet room to manually switch gases and variability in participant ventilation resulting in variability in the respiratory stimulus across participants^7^. At the other end is a system like the RespirAct (Thornhill Medical, Toronto, Canada), which uses four premixed gas sources with different fractions of CO_2_ and O_2_ and computer-controlled flow controllers to deliver precisely controlled gas mixtures. These mixtures are determined by a prospective algorithm to target specific end tidal CO_2_ and/or O_2_ levels^8^, and hence do not rely on measuring expired gases. This reduces variability in the end tidal CO_2_/O_2_ achieved across participants and allows for flexible stimulus design, though it comes with added complexity and higher cost.

This analysis identifies a space for a gas delivery device that automates the switching of gases with predetermined timings, but without the higher cost of more accomplished systems. Therefore, in this study, by taking advantage of low-cost microcontrollers and a simple custom electronic circuit, we developed an automated gas delivery system that can switch between source gases using solenoid valves. The application of this device is demonstrated for a hypercapnic stimulus (elevated carbon dioxide level) and a hyperoxic stimulus (elevated oxygen level) by measuring the direct effect on expired O_2_ and CO_2_ and the resulting physiological response via the BOLD signal.

## Methods

### Gas Delivery Hardware

Gas flow is controlled (Fig 1a) by three solenoid valves from ODE (Colico, Italy): one normally open (21A2ZV30-OX) and two normally closed (21A2KV30-OX), each equipped with a 24 V DC driver (BDA08024CS). At each end of the solenoid valves a male 1/4" BSPP to 8 mm push-fit connector (KQ2H08-02A, SMC, Tokyo, Japan) was fitted and connected via 8 mm pneumatic hosing to a male 1/8” BSPP to 8 mm push-fit connector (KQ2E08-03A). Via 3/8” BSPP to 3/8” BSPP connectors, the output of the solenoid valve was connected to a medical flow meter (Rs, flow-meter, Levate, Italy) and the input was connected to premixed source gases in gas cylinders using a single-stage 4 bar output gas regulator (SSO4, Gas-Arc Group Ltd., Diss, UK). The *normally open* valve was connected to a medical air source to ensure gas flows to the participant even if power to the device is disconnected. However, a *normally closed* valve may be used when the breathing circuit entrains sufficient room air in the absence of a medical air source, as is the case with the short open-ended reservoir used here.

**Figure 1.**
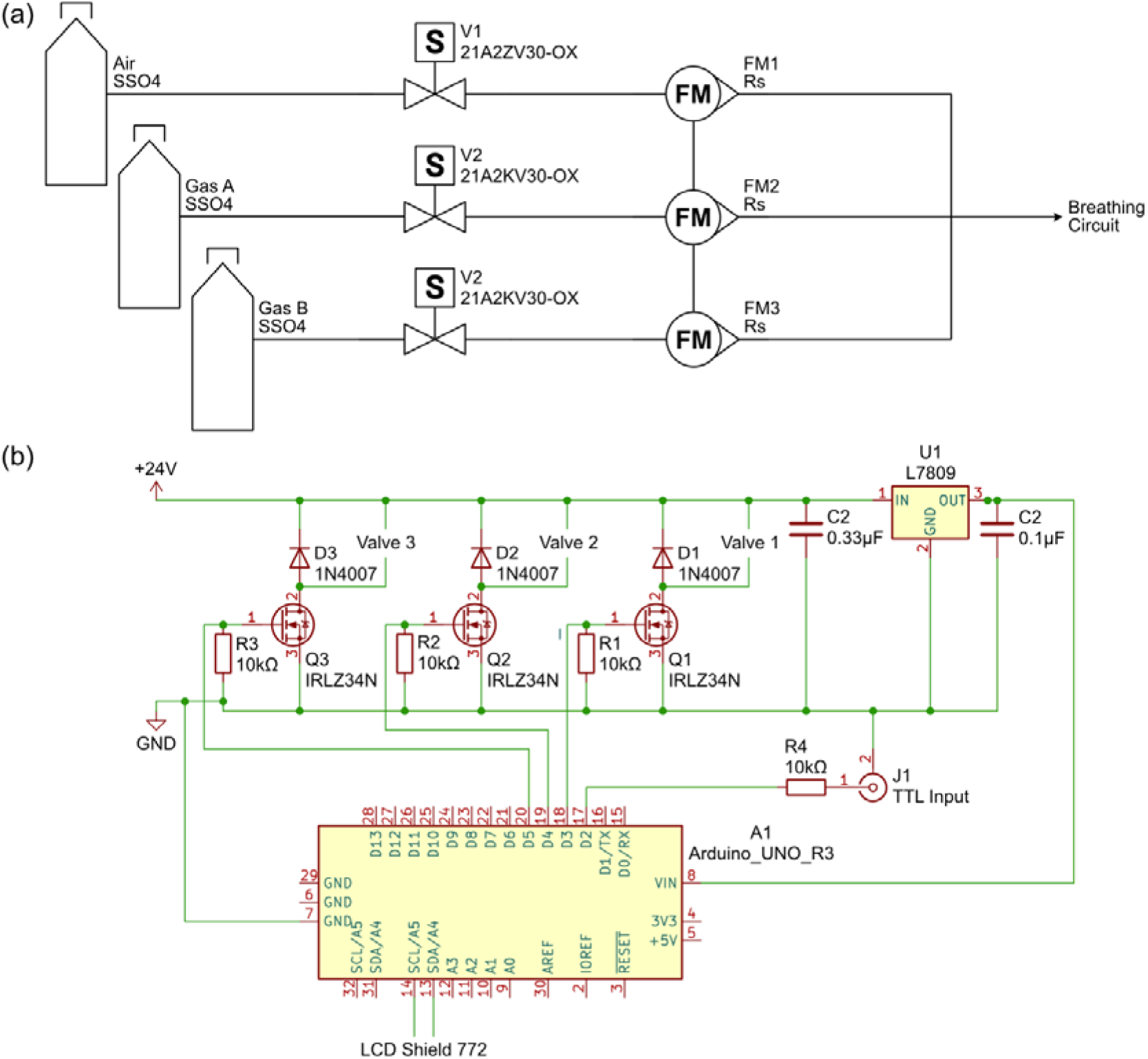
Schematic of the custom controller circuit for the solenoid valves. Each of the three valve channels was driven by an N-channel logic-level MOSFET (Q1–Q3) with a 10LJkΩ pulldown resistor (R1-R3) and 1N4007 flyback diode (D1–D3). The transistor-transistor logic (TTL) input (J1) enables scanner synchronisation and was connected to the Arduino using a 10LJkΩ resistor (R4). A L7809 voltage regulator (U1) and accompanying capacitors (C1, C2) enabled the Arduino, valves and circuit to be driven by the same 24 V power supply.

The control circuitry was based on an Arduino Uno Rev 3 (Arduino, Monza, Italy) and a commercially available LCD shield (772, Adafruit, Brooklyn, NY) with a custom controller circuit for the solenoid valves implemented on a Proto Shield Rev 3 (Arduino, Monza, Italy) sandwiched in between (Fig. 1b). This controller circuit consisted of three N-channel logic-level power MOSFETs (2SK3703-1E, Onsemi, Scottsdale, AZ), one per solenoid valve channel, each connected to a 10 kΩ pulldown resistor and a type 1N4007 flyback diode. Each valve was connected to this circuit using a pre-built cable assembly with built-in back EMF suppression (7000-40881-6360060, Murrelektronik Ltd., Oppenweiler, Germany). A transistor-transistor logic (TTL) input was provided to enable the gas control protocol to be triggered by the TTL output of the MRI scanner. This was connected via a 10 kΩ series resistor to prevent issues with the operation of other apparatus connected to the same TTL signal source. Finally, a voltage regulator (L78S09CV, STMicroelectronics, Geneva, Switzerland), with the recommended 0.1μF and 0.33μF bypass capacitors, was included on the board to step-down the 24 V DC power supply used to energise the solenoid valves to 9 V to power the Arduino microcontroller. Control software was developed using the Arduino IDE to provide three different control modes: (i) a manual mode to control valves manually, (ii) a timer mode triggered by the researcher and (iii) an MRI mode triggered by the scanner TTL output.

Two different embodiments of the gas delivery system were constructed. Firstly, a permanently installed system attached to a rail within the scanner control room with source gases sited outside (Fig. 2a-b). Secondly, a portable system was built and housed within a custom laser cut acrylic case (Fig. 2d-e). At the time of construction, the parts required to build the device in Fig. 2d-e cost ∼£650 GBP (∼$880 USD): 43% solenoid valves, 43% gas fittings including flowmeters, 14% electronics.

**Figure 2.**
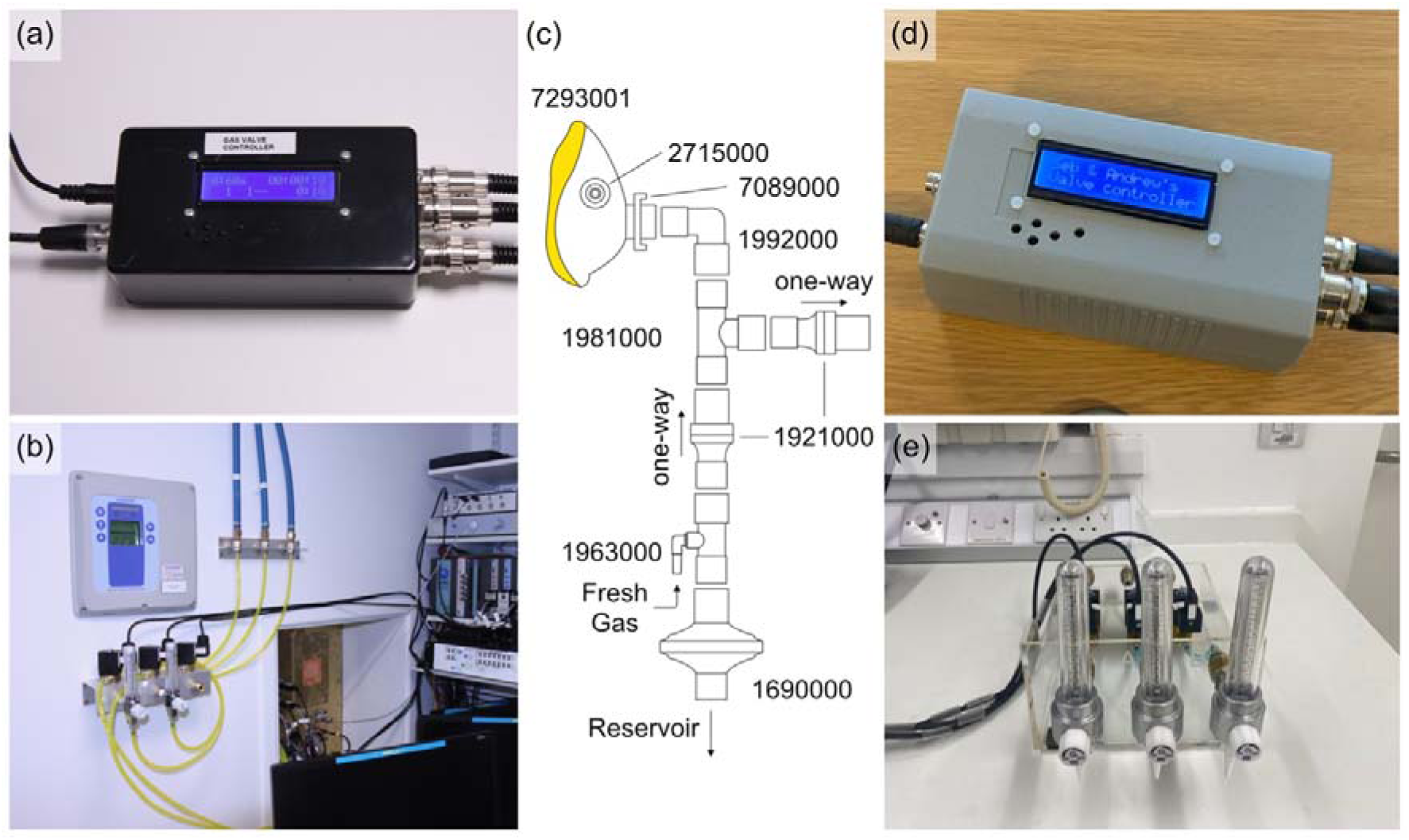
Hardware developed in this study. Two different versions of the gas delivery system were constructed: (b) a system permanently mounted in the MRI scanner control room and (d) a portable system that can be moved between MRI systems. In both cases, (a,d) the microcontroller interface was packaged in a plastic box from which the solenoid valves were energised. Gases from the delivery box were administered to the participant using (c) a breathing circuit consisting of separate inspiratory and expiratory limbs and connected to a reservoir via a breathing filter.

### Participants

Written informed consent was obtained from healthy volunteers following local ethics committee approval (No: FMHS 246-1902). For the hypercapnia experiments, 15 healthy participants (7 male, 8 female, mean age 23±3 years) were included after one exclusion due to an incorrectly set flowmeter delivering insufficient CO_2_. A separate cohort of 15 participants (7 male, 8 female, mean age 27±5 years) was recruited for the hyperoxia experiment.

### Respiratory Stimuli

The outputs of the three medical flow meters were combined using oxygen hosing and two T-pieces and subsequently connected to the breathing circuit via the waveguide into the magnet room. No humidification was used. The breathing circuit (Fig. 2c) is a derivative of a previously described circuit^9^, which uses off the shelf single-use components (Intersurgical, Wokingham, UK), to reduce dead-space volume (changes detailed below). Participants breathed through an anaesthetic mask secured behind their head using a harness which was connected via a fixed elbow (replacing the fixed elbow with sampling port) to a T-piece connector. This formed the inspiratory and expiratory limbs, with direction enforced by one-way valves. The inspiratory limb was connected to a 6 mm stem straight connector, through which fresh gas was delivered through oxygen hosing, and then an inline breathing filter, which enabled a reusable gas reservoir to be connected. The reservoir was constructed of a short 400 mm piece of cuffed tubing, followed by four 2 m long pieces of tubing of the same tubing connected in parallel using three Y-pieces. Whilst this provided a modest reservoir capacity (∼1 litre) its open-ended nature provides improved safety in the event of gas failure. Part numbers are listed in Table 1 and labelled in Fig. 2c (circuit cost ∼£15 GBP/∼$20 USD).

**Table 1.**
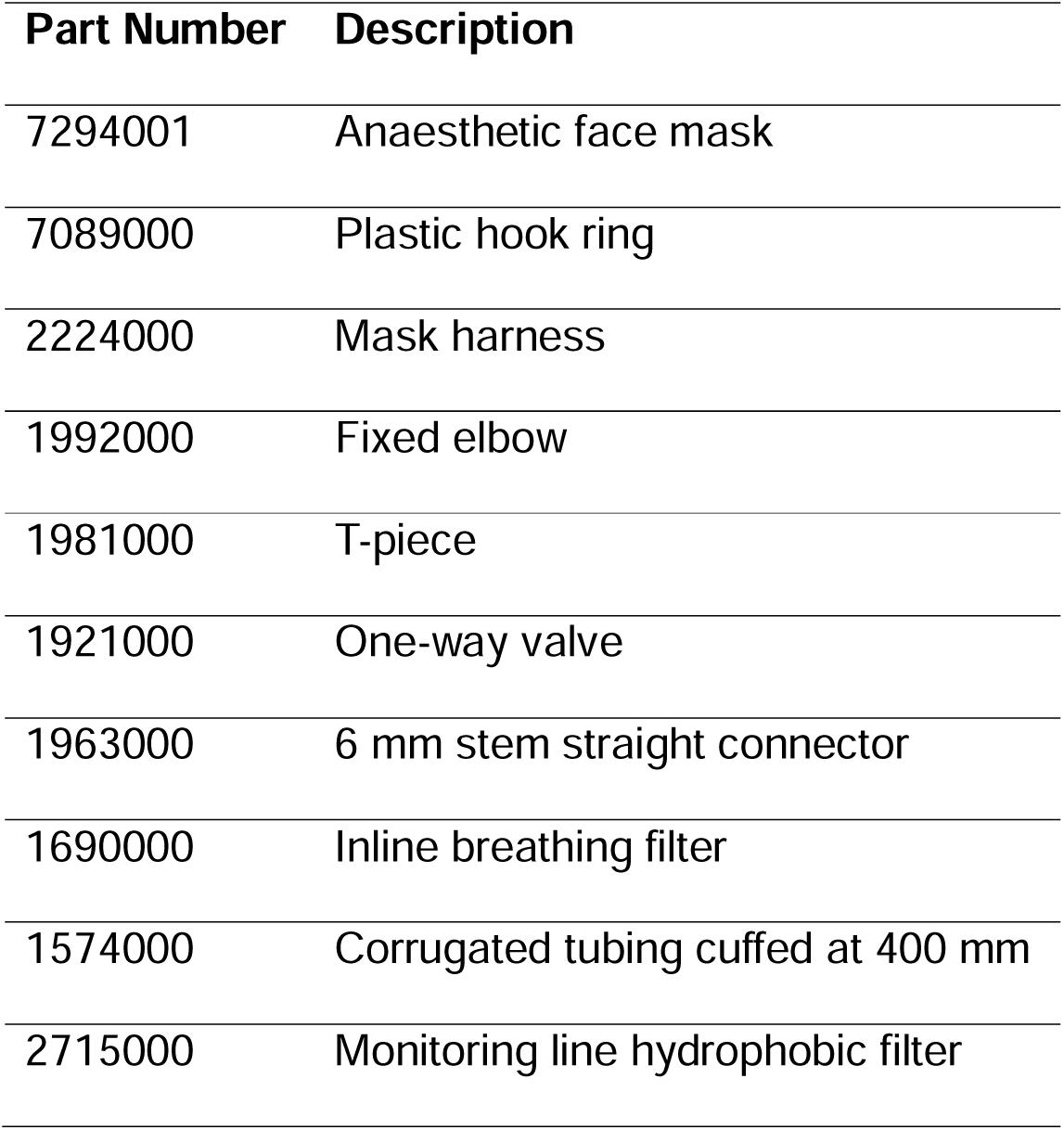
Single use parts for the breathing circuit (Intersurgical, Wokingham, UK). In addition, a female Luer connector with a 1/16” hose barb adapter (45502-00, Masterflex, Gelsenkirchen, Germany) was inserted into the anaesthetic mask in order to sample expired gases.

For expired gas sampling, a female Luer connector with a 1/16” hose barb adapter (45502-00, Masterflex, Gelsenkirchen, Germany) was inserted into the anaesthetic mask (to replace the function of the fixed elbow with sampling port) followed by a hydrophobic filter and a sample line. Expired gases were measured (O_2_ and CO_2_) using a respiratory gas analyser (ML206, AD Instruments, Dunedin, New Zealand). For the hyperoxic stimulus, measurements of CO_2_ were not required for the MRI aspect of the study and hence sensor calibration was not performed although measurements are expected to be linearly proportional to PETCO_2_.

Two respiratory stimuli were implemented: a hypercapnic stimulus and a hyperoxic stimulus. The hypercapnic stimulus consisted of three 60 s blocks of hypercapnia (hypercapnic gas: 5% CO_2_, 21% O_2_, 74% N_2_) interleaved with four 60 s blocks of normocapnia (medical air: 21% O_2_, 79% N_2_) to give a total duration of 7 minutes. The hyperoxia stimulus had two repeats of the following block: 120 s normoxia (medical air: 21% O_2_, 79% N_2_), 30 s hyperoxia (hyperoxic gas: 100% O_2_) and 90 s with both normoxic and hyperoxic gases flowing simultaneously (equivalent to 60.5% O_2_, 39.5% N_2_). This block was repeated twice and followed by a further 120 s normoxic period to give a total duration of 10 minutes. The initial 30 s 100% oxygen block was implemented to produce a large oxygen gradient between the alveolar gas and the blood and results in a rapid increase in end tidal partial pressure of oxygen (PETO_2_). The switch to 60.5% oxygen then rapidly reaches a plateau for the remainder of the block. Each of the medical flow meters were set to 15 lpm which resulted in a total flow of 30 lpm during the plateau of the hyperoxia stimulus.

### Imaging

Data were acquired as part of ongoing imaging studies. In both cases the scanner TTL output was used to trigger the start of the protocol. The hypercapnia stimulus imaging data were acquired on a 3T Premier (GE Healthcare, Waukesha, WI) using multiband EPI (FOV=212×212 mm, matrix 106×106, sixty 2 mm slices, TR/TE=1.4 s/35 ms, multiband factor 3, in-plane parallel acceleration 2, 7 mins duration). The hyperoxia stimulus imaging data were acquired on a 3T Ingenia (Philips Healthcare, Best, NL) using EPI (FOV=224×224 mm, matrix 112×112, thirty 2 mm slices, TR/TE=3 s/30 ms, in-plane parallel acceleration 2, 10 mins duration). In both cases a T_1_-weighted anatomical image was acquired for registration.

### Analysis

Respiratory data were analysed in MATLAB (The Mathworks, Natick, MA). End-tidal timepoints were identified from the peaks of the CO_2_ trace for the hyperoxia stimulus and the troughs of the O_2_ trace for the hypercapnia stimulus. This is useful during transitions in the stimuli as the inspired CO_2_ level can exceed the expired CO_2_ level during CO_2_ increases and likewise the inspired O_2_ level can be lower than the expired O_2_ level during O_2_ decreases. PETO_2_ and end tidal partial pressure of CO_2_ (PETCO_2_) were interpolated onto a 5 s temporal resolution to enable averaging across participants. Group mean values for the baseline and change in PETCO_2_ and PETO_2_ were calculated using time windows. For the hypercapnia stimulus the windows for the baseline were 20 to 30 s, 140 to 150 s and 260 to 270 s and for the peak were 80 to 90 s, 200 to 210 s and 320 to 330 s. Due to a delay in the PETO_2_ response, 30 s were added to these windows when estimating baseline and change in PETO_2_. Whilst for the hyperoxia stimulus the window for the baseline was 0 to 60 s and the windows for the peak were 200 to 230 s and 440 to 470 s.

Imaging data were analysed as follows. Brain extraction was performed using deepbet^10^ on the T_1_-weighted and BOLD images. The BOLD images were motion corrected and subsequently co-registered with the T_1_-weighted images and the MNI atlas^11^ using FMRIB’s Linear Image Registration Tool (FLIRT)^12^. Cortical grey matter regions of interest (ROIs) were created by segmenting the T_1_-weighted images using FMRIB’s Automated Segmentation Tool (FAST)^13^ and then constraining the resulting grey matter partial volume maps to the frontal, insula, occipital, parietal and temporal lobes of the MNI structural atlas. This was then transformed into the BOLD space, thresholded at 50% and used to extract mean BOLD timecourses for each participant. The group mean BOLD signal change was then calculated using the same windows as the respiratory data, but with a 30 s delay relative to the MRI data.

## Results

### Hypercapnia stimulus

Figure 3a shows the changes in PETCO_2_, PETO_2_ and BOLD signal in response to the stimulus with error bars showing the standard error across participants. The group mean change in PETCO_2_ was 8.7±1.8 mmHg (mean±standard deviation) from a baseline of 32.2±3.1 mmHg. The group mean change in PETO_2_ was 10.0±4.5 mmHg from a baseline of 115.6±4.1 mmHg, a percentage change of 8.7±4.0%. The group mean BOLD signal change was 3.2±1.7%, or 0.37%/mmHg when normalised by the change in PETCO_2_.

**Figure 3.**
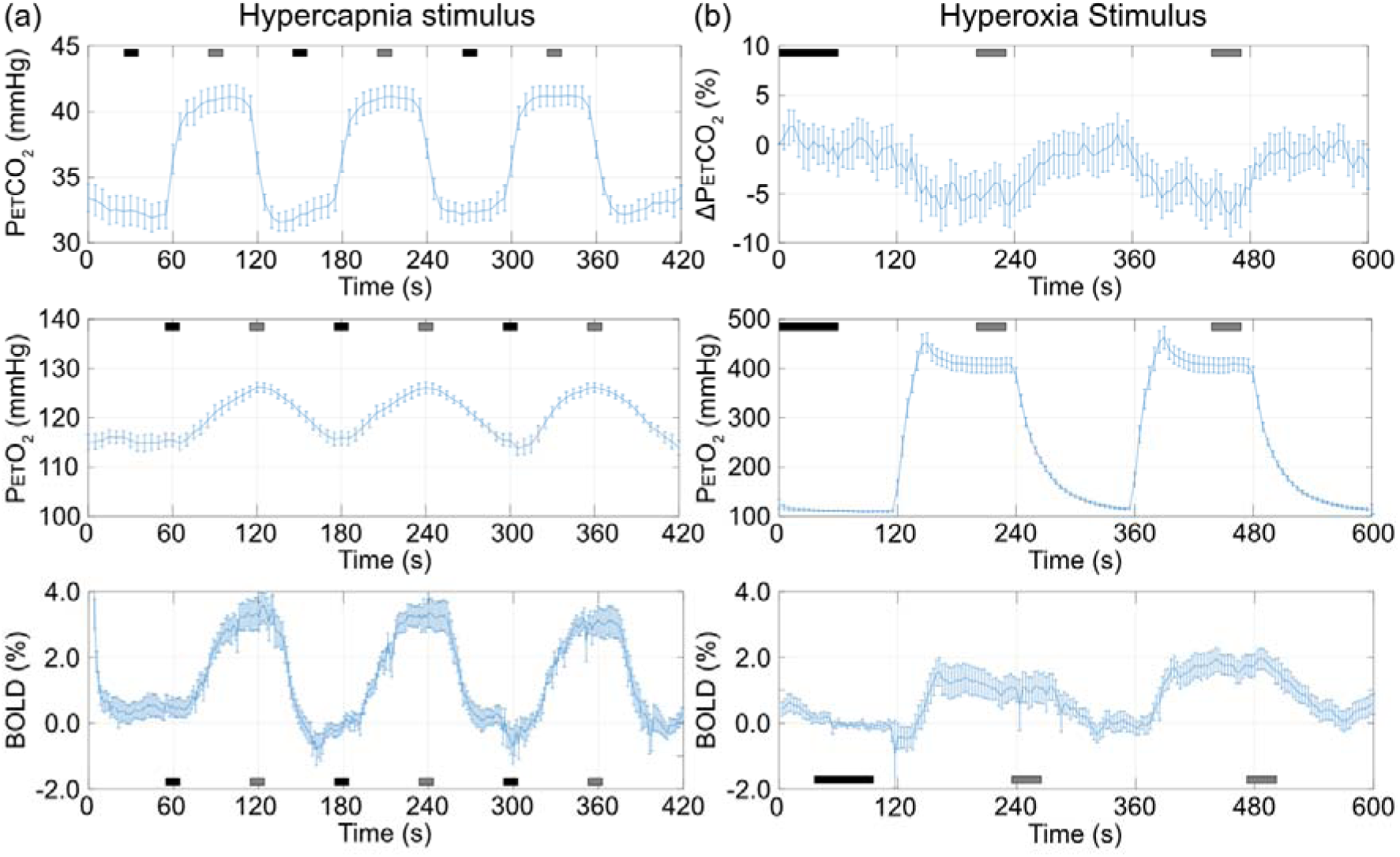
Group mean changes in end-tidal partial pressure of carbon dioxide (PETCO_2_) and oxygen (PETO_2_), or the percentage change in PETCO_2_ (Δ PETCO_2_), in response to (a) a hypercapnic stimulus (5% CO_2_) and (b) a hyperoxic stimulus (initially 100% O_2_, steady state 60.5% O_2_). Error bars represent the standard deviation of the end-tidal partial pressure across the group. Black bars represent the windows for the baseline measurements and grey bars represent the windows for the stimulus measurements. Note: the windows for the BOLD signal include an additional 30 s delay to account for gas delivery, gas sampling and haemodynamic delays.

### Hyperoxia stimulus

Figure 3b shows the changes in PETCO_2_ and PETO_2_ with error bars showing the standard error across participants. The group mean change in PETO_2_ was 265.5±51.3 mmHg from a baseline of 113.2±8.1 mmHg. The group mean percentage change in PETCO_2_ (ΔPETCO_2_) was -4.9±3.8%. The group mean BOLD signal change was 1.2±1.5%, 0.004%/mmHg when normalised by the change in PETO_2_.

## Discussion

This study presents a low-cost, microcontroller-based gas delivery system designed to automate respiratory stimuli for MRI studies. Using affordable components and open-source microcontroller platforms, we demonstrated a flexible and easy to use system for research applications.

### Comparison with Existing Systems

Compared to manual Douglas bag systems, our system improves reproducibility and safety by automating gas switching synchronised with the MRI scanner. Commercial systems like the RespirAct offer end-tidal targeting albeit at higher cost and complexity. In contrast, our system delivers fixed inspired gas mixtures with minimal setup, making it well suited for FIC studies.

Experiments using a similar breathing circuit to this study^14^, demonstrate a slow rise of PETO_2_ in response to a hyperoxia stimulus. With an inspired oxygen fraction (FIO_2_) of 50% a plateau was reached in 3 minutes. In contrast, we demonstrate that by using a short period with an FIO_2_ of 100% a rapid increase in PETO_2_ is achievable due to the large gradient between alveolar and blood PO_2_. A plateau at an FIO_2_ of 60.5% can then be reached within 30 s.

BOLD measurements provided physiological validation of the respiratory stimuli. The BOLD response to hypercapnia is typically reported as the percentage signal change per mmHg change in PETCO_2_ for grey matter. The value of 0.37%/mmHg measured in this study is in line with values (0.35%/mmHg) reported in a recent systematic review^15^. Whilst the BOLD response to hyperoxia in grey matter isn’t commonly reported in this way, the measured value of 0.004%/mmHg falls within the range (0.006%/mmHg) of a previous study^16^. Although qualitative in nature, these results demonstrate that the system is well suited to applications such as CVR mapping^17^ and hyperoxiaLJBOLD CBV estimation^18^.

### Strengths

The system has several strengths. First, the system is cost effective with a total build cost that is significantly lower than commercial alternatives. Secondly, the Arduino-based microcontrollers and off the shelf components makes the system easy to replicate and modify. Thirdly, the design of the system is open source with hardware schematics and control software shared freely. Finally, the system operates outside the scanner room with gases routed into the shielded room, ensuring magnet safety.

### Limitations

Despite its strengths, the system has some limitations. The system cannot dynamically blend gases and hence it is not able to implement dynamic end-tidal forcing or prospective end-tidal targeting algorithms to more precisely control PETCO_2_ and PETO_2_. Therefore, the system only supports block design stimuli, which may not suit all paradigms. It can also be seen that modulating one respiratory gas has a concomitant effect on the other, which may be mitigated by clamping the latter using a sequential gas delivery (SGD) circuit^19^. Whilst, as noted above, the absolute PETCO_2_ change during the hyperoxia stimulus cannot be relied upon because of a lack of calibration, the percentage change is small compared to the hypercapnia stimulus (∼6% reduction vs ∼27% increase). Likewise, the change in PETO_2_ is also small for the hypercapnia stimulus when compared to the hyperoxia stimulus (∼0.5% reduction vs ∼255%). Finally, whilst systems such as this can be used in a research context, they are not approved for clinical use.

### Future Work

Several enhancements could improve the system. With an appropriate breathing circuit, sequential gas delivery with a rebreathe bag could be implemented^19^ to clamp PETO_2_ or PETCO_2_ for hyperoxic or hypercapnic stimuli, respectively. This would reduce variability caused by increased ventilation rate, although it would not improve intersubject variability.

From a software perspective, the current implementation can only save 10 hard coded paradigms. For developmental studies it would be useful to expand the user interface’s functionality to allow editing and saving custom paradigms or to add a host-client mode allowing an external laptop to control the microcontroller, allowing greater flexibility in stimulus design.

## Conclusion

In this study we have demonstrated a low-cost, microcontroller-based gas delivery system for administering fixed inspired gas challenges. The use of solenoid valves enables reliable automation without the expense of flow controllers and hence sits in a gap between manual and commercial gas delivery systems. Its affordability and ease of use could make it particularly attractive for research groups with limited access to commercial gas delivery systems.

## Data Availability Statement

Hardware schematics and Arduino valve controller code are made available at: https://github.com/fmriphysiology/mri-gas-delivery-system/ (release v1.0.0). MATLAB scripts are available at: https://github.com/fmriphysiology/gasdelivery (release v1.0.0). Expired gases data are available at, doi: https://doi.org/10.5281/zenodo.19097161.

## Acknowledgements

The authors thank Stuart Salter and Ian Taylor for their invaluable technical assistance with the mechanical and electronics aspects of this study, respectively. This research was funded by the Nottingham BBSRC Doctoral Training Partnership, a PhD studentship from Taif University, the EPSRC (EP/S021507/1) and supported by the NIHR Nottingham Biomedical Research Centre (NIHR203310) and the NIHR Oxford Health Biomedical Research Centre (NIHR203316). The views expressed are those of the author(s) and not necessarily those of the NIHR or the Department of Health and Social Care. The Centre for Integrative Neuroimaging was supported by core funding from the Wellcome Trust (203139/Z/16/Z and 203139/A/16/Z). This research was funded in part by UKRI. For the purpose of Open Access, the author has applied a CC BY public copyright licence to any Author Accepted Manuscript version arising from this submission.

